# Risk factors for heart failure with preserved or reduced ejection fraction among Medicare beneficiaries: Applications of competing risks analysis and gradient boosted model

**DOI:** 10.1101/19003509

**Authors:** Moa P. Lee, Robert J. Glynn, Sebastian Schneeweiss, Kueiyu Joshua Lin, Elisabetta Patorno, Julie Barberio, Raisa Levin, Thomas Evers, Shirley V. Wang, Rishi J. Desai

**Affiliations:** Division of Pharmacoepidemiology and Pharmacoeconomics, Brigham and Women’s Hospital & Harvard Medical School, Boston, MA, USA; Department of Epidemiology, University of North Carolina, Chapel Hill, NC, USA; Department of Medicine, Massachusetts General Hospital & Harvard Medical School, Boston, MA, USA; Bayer AG, Wuppertal, Germany

**Author notes:** Correspondence: Rishi J Desai, MS, PhD, Division of Pharmacoepidemiology and Pharmacoeconomics, Department of Medicine, Brigham and Women’s Hospital and Harvard Medical School, 1620 Tremont Street, Boston, MA 02120, USA, |.

**Keywords:** heart failure, epidemiology, risk factors, LVEF, HEpEF, HFrEF, competing risks, GBM

## Abstract

**Background:** The differential impact of various demographic characteristics and comorbid conditions on development of heart failure (HF) with preserved (pEF) and reduced ejection fraction (rEF) is not well studied among the elderly.

**Methods and Results:** Using Medicare claims data linked to electronic health records, we conducted an observational cohort study of individuals ≥ 65 years of age without HF. A Cox proportional hazards model accounting for competing risk of HFrEF and HFpEF incidence was constructed. A gradient boosted model (GBM) assessed the relative influence (RI) of each predictor in development of HFrEF and HFpEF. Among 138,388 included individuals, 9,701 developed HF (IR= 20.9 per 1,000 person-year). Males were more likely to develop HFrEF than HFpEF (HR = 2.07, 95% CI: 1.81-2.37 vs. 1.11, 95% CI: 1.02-1.20, *P* for heterogeneity < 0.01). Atrial fibrillation and pulmonary hypertension had stronger associations with the risk of HFpEF (HR = 2.02, 95% CI: 1.80-2.26 and 1.66, 95% CI: 1.23-2.22) while cardiomyopathy and myocardial infarction were more strongly associated with HFrEF (HR = 4.37, 95% CI: 3.21-5.97 and 1.94, 95% CI: 1.23-3.07). Age was the strongest predictor across all HF subtypes with RI from GBM >35%. Atrial fibrillation was the most influential comorbidity for development of HFpEF (RI = 8.4%) while cardiomyopathy was most influential for HFrEF (RI = 20.7%).

**Conclusions:** These findings of heterogeneous relationships between several important risk factors and heart failure types underline the potential differences in the etiology of HFpEF and HFrEF.

**Key Questions:**

- What is already known about this subject? Previous epidemiologic studies describe the differences in risk factors involved in developing heart failure with preserved (HFpEF) and reduced ejection fraction (HFrEF), however, there has been no large study in an elderly population.
- What does this study add? This study provides further insights into the heterogeneous impact of various clinical characteristics on the risk of developing HFpEF and HFrEF in a population of elderly individuals. Employing an advanced machine learning technique allows assessing the relative importance of each risk factor on development of HFpEF and HFrEF.
- How might this impact on clinical practice? Our findings provide further insights into the potential differences in the etiology of HFpEF and HFrEF, which are critical in prioritizing populations for close monitoring and targeting prevention efforts.

## INTRODUCTION

Heart failure is a highly debilitating disease affecting 6.5 million individuals in the US. (1) The incidence of heart failure rises substantially with age, and it is estimated that heart failure accounts for approximately 25% of all hospitalizations in patients over 65 years old. (2-4) Heart failure is classified into two major subtypes based on systolic function measured by left ventricular ejection fraction (LVEF), 1) heart failure with preserved ejection fraction (HFpEF), and 2) heart failure with reduced ejection fraction (HFrEF). The pathophysiology of HFrEF is well-understood and involves left ventricular eccentric hypertrophy and markedly reduced end-systolic elastance. (5) However, HFpEF is a complex syndrome with an evolving understanding of its pathophysiology and involves abnormalities in left ventricular relaxation and compliance, which may result from systemic proinflammatory states induced by salient co-morbid conditions including hypertension, obesity, diabetes, and chronic obstructive pulmonary disease. (6, 7)

Risk factors for heart failure identified as a composite of HFpEF and HFrEF, are well-described in the literature and include coronary disease, particularly myocardial infarction and valvular disease, renal disease, diabetes, and obesity. (2, 3, 8, 9) However, only a few investigations have focused on differentiating factors that may have heterogeneous associations with HFpEF and HFrEF. (10-12) As many established treatments of heart failure, including inhibitors of the renin-angiotensin system and beta-blockers provide clear benefit in HFrEF but little benefit in HFpEF patients, (13, 14) there has been a revived interest in understanding etiologies and phenotypes that may differentiate heart failure with pEF from rEF. (15) Since presentation of heart failure has changed over time with a growing incidence but no improvement in outcomes in HFpEF, (16-18) it is critical to validate findings regarding differences in risk factors leading to HFpEF and HFrEF observed in previous studies.(10, 11, 19) Thus, in this study, we sought to compare risk factors for HFpEF and HFrEF in a large population of Medicare beneficiaries using a competing risk analysis, which offers the convenience of handling these factors simultaneously while allowing the flexibility of examining the associations with each type of outcome separately. (20)

## METHODS

### Data sources and study cohort

We conducted a cohort study using 2007-2014 Medicare claims data from Parts A (inpatient coverage), B (outpatient coverage), and D (prescription benefits) for this study. Medicare claims were linked deterministically by health insurance claim (HIC) numbers, date of birth, and sex with electronic health records (EHR) for two large healthcare provider networks in the Boston metropolitan area. The study protocol was approved by the Institutional Review Board of the Brigham and Women’s Hospital, and patient informed consent was not required as the database was de-identified to protect subject confidentiality.

Patients aged 65 or above with at least six months of continuous enrollment in Medicare (parts A, B, and D) were identified between 2007 and 2014 for inclusion in this study. The date when these patients met this requirement was considered the cohort entry date and the 6-month continuous enrollment period prior to the cohort entry date was defined as the baseline period. To ensure that the patients were free of heart failure at the time of cohort entry, patients were required to have no diagnosis, inpatient or outpatient, of heart failure, as defined in **Supplementary Table 1**, during the baseline period. We further excluded patients using heart failure medications including mineralocorticoid receptor antagonist (eplerenone, spironolactone), digoxin, or loop diuretics (bumetanide, furosemide, torsemide, ethacrynic acid) during baseline. To minimize the degree of missingness due to discontinuity of care in the EHR system, we further limited our study cohort to patients with at least one ICD-9 diagnosis in EHRs any time during the study period. (21) Follow-up for occurrence of heart failure started on the cohort entry date and ended on the first occurrence of any of the following censoring events: disenrollment from Medicare, death, or the end of follow-up.

### Identification and classification of heart failure

The first inpatient diagnosis of heart failure from Medicare claims using ICD-9 codes (see **Supplementary Table 1**) was identified during follow-up. We classified these heart failure cases as HFrEF, HFpEF or uncertain EF by extracting LVEF values that were most proximal to the date of heart failure diagnosis from electronically recorded cardiology reports (echocardiograms or cardiac catheterization) using natural language processing. Specifically, the Medical Text Extraction, Reasoning and Extraction System (22) was used to extract expressions identifying LVEF along with important contextual information such as negations in reports and collect the nearest valid value to the expression. A sample of the notes (n=200) was manually reviewed to ensure the basic rules of LVEF extraction were working. Rules were further refined by manually reviewing all false positives for these 200 notes and removing sources of obvious errors. We then validated a random sample 100 of notes by comparing NLP extracted EF values against manual extraction by a chart reviewer. In 99 of these notes, the NLP algorithm extracted numerical EF values accurately. At this point, the NLP algorithm was deemed reliable and was used for extraction of EF values from notes of all included patients. Based on EF values recorded around the time of HF diagnosis, within 1 month prior to the first episode of heart failure up to 1 year after the diagnosis, the cases were classified into heart failure with reduced EF defined as LVEF <45% or with preserved EF defined as LVEF ≥ 45%. (23) Patients with heart failure diagnosis without any evidence of LVEF recordings around the time of the diagnosis in our EHR were still included in the analysis and classified to be heart failure with uncertain EF.

### Assessment of risk factors

Several patient characteristics were considered as potential risk factors based on their reported role in development of heart failure in prior studies. These risk factors were assessed during the baseline period prior to the cohort entry date and included demographic and lifestyle factors, such as age, gender, race, smoking, as well as comorbidities such as atrial fibrillation, anemia, cardiomyopathy, chronic obstructive pulmonary disease (COPD), depression, diabetes, hyperkalemia, hypertension, myocardial infarction, obesity, valvular heart disease, and chronic renal disease (see **Supplementary Table 2** for the complete list of antecedent patient factors). (8-10, 24) Patient demographics were identified from enrollment files and comorbidities were identified by the occurrence of diagnosis codes (**Supplementary Table 2**) in Medicare inpatient or outpatient claims.

### Statistical Analysis

In descriptive analyses, we examined the distributions of the risk factors among the study cohort. We calculated the incidence rate of heart failure in four age groups (65-69, 70-74, 75-79 and ≥ 80 years). A standard Cox proportional hazards model was used to assess the effects of each risk factor on incident heart failure. To estimate the associations of each risk factor with the relative hazard of each heart failure type, we used an extension of the Cox proportional hazards model accounting for competing risk through the methods of data augmentation proposed by Lunn and McNeil. (20, 25) Data augmentation involved duplicating the data 3 times for each heart failure type allowing each subject to have a separate observation for each outcome. Using the augmented data, Kaplan-Meier curves was constructed for each outcome to assess the cumulative hazard function for each failure type. Then we fitted a Cox regression model, stratified by heart failure type, including the interaction terms between each risk factor and heart failure type allowing different associations of each risk factor with each specific heart failure type. We assessed and reported the P values for the interaction terms for the tests that the effects of the risk factors were the same across the outcomes.

To provide further insight into the potential differences in the relationships between individual risk factors and each heart failure type, a gradient boosted model (GBM) adapted for the Cox model was applied. (26, 27) The importance of each risk factor on outcomes was assessed based on the estimated variable influence from models fitted separately for each type of heart failure. The relative influence of each variable is calculated based on the improvement in reducing the loss function – analogous to the deviance in logistic regression – from splitting the regression tree using a predictor and how often the predictor is selected for the split, (26, 27) reflecting the relative importance of the variable in improving the predictive ability of the model. For GBM, we specified 10,000 boosting iterations, a learning rate parameter of 0.01 and an interaction depth of 1 for the additive approximations. These specifications were chosen because a small learning rate parameter when combined with a large number of iterations is known to reduce prediction error in GBM. (28) The overall performance of the models were assessed by the C index proposed by Pencina et al. (29, 30) All statistical analyses were performed using SAS 9.4 (SAS Institute, Cary NC) and R 3.2.0 (R Foundation for Statistical Computing, Vienna, Austria).

## RESULTS

A total of 138,388 Medicare beneficiaries were included in the analysis (**Figure 1**); the mean age of the cohort was 71.9 ± 6.9 with 38% male (**Table 1**). Hyperlipidemia and hypertension were the most prevalent comorbidities affecting over 50% of the study cohort followed by psychosis appearing in 27% of the study cohort. Diabetes and anemia affected 18% and 13% of the study cohort, respectively.

**Table 1.** Baseline Characteristics of the Study Cohort (n=138,388)

| Table 1. Baseline Characteristics of the Study Cohort (n=138,388) |  |  |
| --- | --- | --- |
|  | No. | % |
| Variables |  |  |
| Demographic |  |  |
| Age - mean (SD) | 71.9 | 6.9 |
| Age categories |  |  |
| 65-69 | 66,712 | 48.2 |
| 70-74 | 28,746 | 20.8 |
| 75-79 | 20,809 | 15.0 |
| 80+ | 22,121 | 16.0 |
| Male | 53,093 | 38.1 |
| Race |  |  |
| White | 124,594 | 90.0 |
| Black | 4,680 | 3.4 |
| Other | 9,114 | 6.6 |
| Comorbidities |  |  |
| Atrial fibrillation | 8,351 | 6.0 |
| Anemia | 18,471 | 13.4 |
| Cardiomyopathy | 1,437 | 1.0 |
| COPD | 9,829 | 7.1 |
| Depression | 15,533 | 11.2 |
| Diabetes | 24,866 | 18.0 |
| Hyperkalemia | 1,251 | 0.9 |
| Hyperlipidemia | 87,149 | 63.0 |
| Hypertension | 76,596 | 55.4 |
| Hypotension | 2,241 | 1.6 |
| Myocardial infarction | 1,034 | 0.8 |
| Obesity | 5,778 | 4.2 |
| Other dysrhythmia | 11,116 | 8.0 |
| Psychosis | 38,065 | 27.3 |
| Pulmonary hypertension | 793 | 0.6 |
| Sleep apnea | 3,380 | 2.4 |
| Smoking | 7,195 | 5.2 |
| Angina | 2,948 | 2.1 |
| Stroke | 4,099 | 3.0 |
| TIA | 1,914 | 1.4 |
| Valvular heart disease | 4,326 | 3.1 |
| Chronic renal disease | 5,314 | 3.8 |
| Renal insufficiency | 1,326 | 1.0 |

COPD = chronic obstructive pulmonary disease; TIA = transient ischemic attack

**Figure 1.**
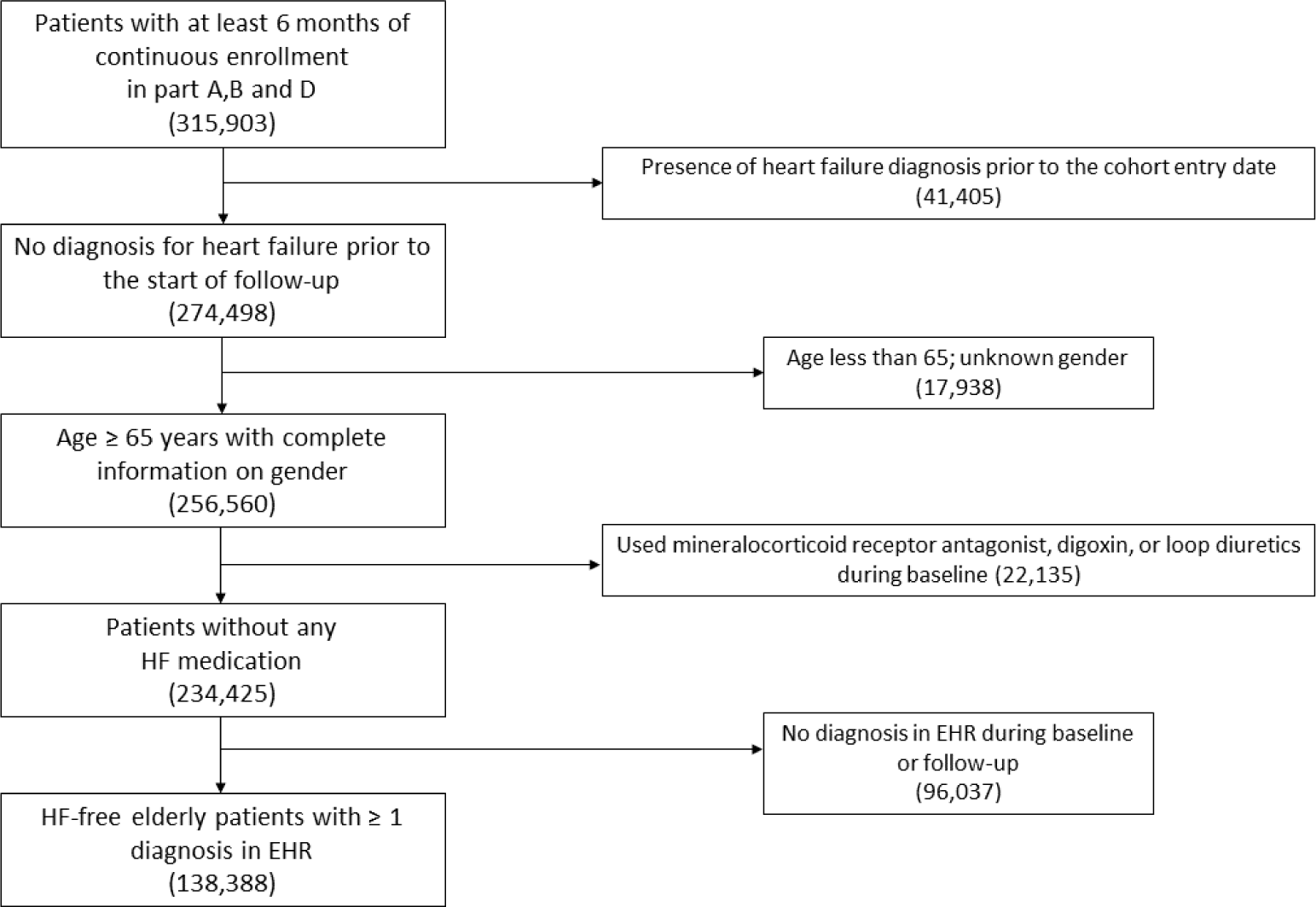
Patient Disposition.

During a mean follow-up of 3.4 ± 1.7 years with a total of 463,710 person-years of follow-up accrued, 9,701 patients developed heart failure. The median (IQR) time to heart failure was 2.3 (1.1 - 3.6) years. Of the 9,701 cases, 2,806 (28.9%) had preserved LVEF, and 923 (9.5%) had reduced LVEF while 5,972 (61.6%) of heart failure cases had no record of LVEF in the linked EHR. The incidence rate of heart failure was 20.9 per 1,000 person-years; the incidence rates of heart failure with preserved, reduced, and uncertain EF were 6.1, 2.0, and 12.9 per 1,000 person-years, respectively. Incidence was higher in men compared to women (23.5 vs. 19.3 per 1,000 person-years) with the rates being substantially higher for the cases of heart failure with reduced EF among men compared to women (2.9 vs. 1.4 per 1,000 person-years). The risk of developing heart failure increased with age, from 11.1 per 1,000 person-years among patients aged 65 to 69 years to 49.2 per 1,000 person-years in patients aged 80 years or greater (**Table 2**).

**Table 2.** Incidence Rates of Heart Failure Among All Cohort, by Gender and Age Group

|  | Heart failure |  | Preserved EF |  | Reduced EF |  | Uncertain EF |  |
| --- | --- | --- | --- | --- | --- | --- | --- | --- |
|  | No. of events | IR*<br>(95% CI) | No. of events | IR*<br>(95% CI) | No. of events | IR*<br>(95% CI) | No. of events | IR*<br>(95% CI) |
| All cohort | 9,701 | 20.9<br>(20.5-21.3) | 2,806 | 6.1<br>(5.8-6.3) | 923 | 2.0<br>(1.9-2.1) | 5,972 | 12.9<br>(12.6-13.2) |
| Gender |  |  |  |  |  |  |  |  |
| Women | 5,485 | 19.3<br>(18.8-19.8) | 1,659 | 5.8<br>(5.6-6.1) | 398 | 1.4<br>(1.3-1.5) | 3,428 | 12.1<br>(11.7-12.5) |
| Men | 4,216 | 23.5<br>(22.8-24.2) | 1,147 | 6.4<br>(6.0-6.8) | 525 | 2.9<br>(2.7-3.2) | 2,544 | 14.2<br>(13.6-14.7) |
| Age group |  |  |  |  |  |  |  |  |
| 65-69 | 2,419 | 11.1<br>(10.7-11.6) | 685 | 3.1<br>(2.9-3.4) | 250 | 1.2<br>(1.0-1.3) | 1,484 | 6.8<br>(6.5-7.2) |
| 70-74 | 1,794 | 17.6<br>(16.8-18.4) | 523 | 5.1<br>(4.7-5.6) | 174 | 1.7<br>(1.5-2.0) | 1,097 | 10.8<br>(10.2-11.4) |
| 75-79 | 2,018 | 27.4<br>(26.2-28.6) | 578 | 7.8<br>(7.2-8.5) | 216 | 2.9<br>(2.6-3.3) | 1,224 | 16.6<br>(15.7-17.5) |
| 80+ | 3,470 | 49.2<br>(47.6-50.9) | 1,020 | 14.5<br>(13.6-15.4) | 283 | 4.0<br>(3.6-4.5) | 2,167 | 30.7<br>(29.5-32.1) |

\*Incidence rates (IR) are in 1,000 person-years

### Associations of clinical characteristics with incident heart failure

Risk of heart failure increased substantially with age (hazard ratio per 5 years of age = 1.46; 95% confidence interval: 1.44, 1.48). Men were more likely to develop heart failure (hazard ratio= 1.22; 95% confidence interval: 1.17, 1.28) compare to women (**Table 3**). Atrial fibrillation and cardiomyopathy were associated with an elevated risk of heart failure with hazard ratio of 1.90 (95% confidence interval: 1.79, 2.02) and 2.22 (95% confidence interval: 1.97, 2.50), respectively. COPD was also positively associated with heart failure with hazard ratio of 1.87 (95% confidence interval: 1.76, 1.98). Diabetes was associated with an increased risk (hazard ratio = 1.72; 95% confidence interval: 1.64, 1.80), whereas being diagnosed with hyperlipidemia was associated with decreased risk (hazard ratio = 0.91; 95% confidence interval: 0.87, 0.95). Obesity and smoking were associated with an increased risk (hazard ratio = 1.15; 95% confidence interval: 1.03, 1.27 and 1.33; 95% confidence interval: 1.22, 1.44, respectively). Other cardiovascular related comorbidities including hypertension, dysrhythmia, valvular heart disease, stroke and angina were associated with increased risk of heart failure. Patients with baseline diagnosis of anemia, hyperkalemia or chronic renal disease were more likely to develop heart failure than those without such comorbidities.

**Table 3.** Associations of Risk Factors With Developing Heart Failure (9,701 cases) Among the Study Cohort

|  | HR<sup>*</sup> | 95% CI |  |
| --- | --- | --- | --- |
| Variables |  |  |  |
| Demographic |  |  |  |
| Age (per 5 years) | 1.46 | 1.44 | 1.48 |
| Gender |  |  |  |
| Female | 1.00 | Referent |  |
| Male | 1.22 | 1.17 | 1.28 |
| Race |  |  |  |
| White | 1.00 | Referent |  |
| Black | 1.01 | 0.90 | 1.12 |
| Other | 0.82 | 0.74 | 0.90 |
| Comorbidities |  |  |  |
| Atrial fibrillation | 1.90 | 1.79 | 2.02 |
| Anemia | 1.27 | 1.21 | 1.34 |
| Cardiomyopathy | 2.22 | 1.97 | 2.50 |
| COPD | 1.87 | 1.76 | 1.98 |
| Depression | 1.02 | 0.95 | 1.09 |
| Diabetes | 1.72 | 1.64 | 1.80 |
| Hyperkalemia | 1.39 | 1.20 | 1.61 |
| Hypertension | 1.18 | 1.13 | 1.24 |
| Hypotension | 0.95 | 0.83 | 1.08 |
| Myocardial infarction | 1.10 | 0.94 | 1.30 |
| Obesity | 1.15 | 1.03 | 1.27 |
| Other dysrhythmia | 1.14 | 1.07 | 1.22 |
| Psychosis | 0.99 | 0.94 | 1.04 |
| Hyperlipidemia | 0.91 | 0.87 | 0.95 |
| Pulmonary hypertension | 1.26 | 1.06 | 1.50 |
| Valvular heart disease | 1.45 | 1.32 | 1.59 |
| Sleep apnea | 1.11 | 0.98 | 1.27 |
| Smoking | 1.33 | 1.22 | 1.44 |
| Stroke | 1.27 | 1.16 | 1.39 |
| TIA | 0.81 | 0.70 | 0.94 |
| Angina | 1.17 | 1.06 | 1.30 |
| Renal insufficiency | 1.15 | 0.98 | 1.36 |
| Chronic renal disease | 1.58 | 1.46 | 1.71 |

\*adjusted for all other risk factors listed in the table
COPD = chronic obstructive pulmonary disease; TIA = transient ischemic attack

### Associations of clinical characteristics with heart failure types

#### Competing risks analysis

Table 4 summarized the associations of risk factors with each heart failure type and whether each risk factor has a different association across the outcomes. While age, diabetes, and chronic renal disease were positively associated with the risk of heart failure across all types (P value for heterogeneity > 0.05), several risk factors had differential relations with the risk of heart failure across the different types. For instance, male gender was associated with an increased the risk for heart failure with rEF (hazard ratio = 2.07; 95% confidence interval: 1.81, 2.37) more substantially than heart failure with pEF. Patients with atrial fibrillation at baseline were more likely to develop heart failure with pEF (hazard ratio = 2.02; 95% confidence interval: 1.80, 2.26) whereas cardiomyopathy was more strongly associated with heart failure with rEF (hazard ratio = 4.37; 95% confidence interval: 3.21, 5.97). Obesity and pulmonary hypertension were more strongly associated with the risk for heart failure with pEF (hazard ratio = 1.42; 95% confidence interval: 1.19, 1.69 and hazard ratio = 1.66; 95% confidence interval: 1.23, 2.22, respectively) while history of myocardial infarction was most strongly associated with heart failure with rEF (hazard ratio = 1.94; 95% confidence interval: 1.23, 3.07).

#### GBM

Variable influence obtained from GBM differed notably across each type of heart failure (**Figure 2**). Age was the strongest predictor of all heart failure types contributing to more than 35% of the model’s predictive ability. Atrial fibrillation was the most determining comorbidity for heart failure with pEF accounting for 8.4% of the outcome prediction. Cardiomyopathy was most predictive of heart failure with rEF (relative influence = 20.7%). Male gender and history of myocardial infarction more strongly determined the risk of heart failure with rEF (relative influence = 11.8% and 5.3%, respectively). Diabetes appeared to be influential in predicting heart failure across all types with the relative influence above 5% in all heart failure types.

**Table 4.** Associations of Risk Factors With Developing Heart Failure With Reduced, Preserved, or Uncertain LVEF

|  | Heart failure with<br>preserved EF*<br>(2,806 cases) |  |  |  | Heart failure with<br>reduced EF†<br>(923 cases) |  |  |  | Heart failure with<br>uncertain EF‡<br>(5,972 cases) |  |  |  |
| --- | --- | --- | --- | --- | --- | --- | --- | --- | --- | --- | --- | --- |
|  | HR | 95% CI |  |  | HR | 95% CI |  |  | HR | 95% CI |  | *P* Heterogeneity |
| Variables |  |  |  |  |  |  |  |  |  |  |  |  |
| Demographic |  |  |  |  |  |  |  |  |  |  |  |  |
| Age (per 5 years) | 1.48 | 1.44 | 1.51 |  | 1.41 | 1.35 | 1.47 |  | 1.46 | 1.44 | 1.49 | 0.22 |
| Gender |  |  |  |  |  |  |  |  |  |  |  |  |
| Female | 1.00 | Referent |  |  | 1.00 | Referent |  |  | 1.00 | Referent |  |  |
| Male | 1.11 | 1.02 | 1.20 |  | 2.07 | 1.81 | 2.37 |  | 1.18 | 1.12 | 1.24 | <0.01 |
| Race |  |  |  |  |  |  |  |  |  |  |  |  |
| White | 1.00 | Referent |  |  | 1.00 | Referent |  |  | 1.00 | Referent |  |  |
| Black | 1.38 | 1.15 | 1.65 |  | 1.47 | 1.08 | 1.99 |  | 0.77 | 0.66 | 0.90 | <0.01 |
| Other | 1.06 | 0.90 | 1.24 |  | 0.91 | 0.68 | 1.21 |  | 0.69 | 0.61 | 0.79 | <0.01 |
| Comorbidities |  |  |  |  |  |  |  |  |  |  |  |  |
| Atrial fibrillation | 2.02 | 1.80 | 2.26 |  | 1.39 | 1.11 | 1.73 |  | 1.93 | 1.79 | 2.08 | 0.01 |
| Anemia | 1.28 | 1.17 | 1.41 |  | 1.02 | 0.85 | 1.23 |  | 1.30 | 1.22 | 1.39 | 0.05 |
| Cardiomyopathy | 1.84 | 1.45 | 2.34 |  | 4.37 | 3.21 | 5.97 |  | 2.12 | 1.82 | 2.47 | <0.01 |
| COPD | 1.58 | 1.40 | 1.78 |  | 1.55 | 1.25 | 1.92 |  | 2.05 | 1.90 | 2.20 | <0.01 |
| Depression | 0.97 | 0.86 | 1.10 |  | 0.73 | 0.57 | 0.95 |  | 1.08 | 0.99 | 1.16 | 0.01 |
| Diabetes | 1.65 | 1.51 | 1.80 |  | 1.67 | 1.43 | 1.94 |  | 1.75 | 1.65 | 1.86 | 0.49 |
| Hyperkalemia | 1.33 | 0.99 | 1.77 |  | 1.39 | 0.84 | 2.31 |  | 1.42 | 1.19 | 1.70 | 0.92 |
| Hypertension | 1.25 | 1.14 | 1.36 |  | 1.03 | 0.89 | 1.20 |  | 1.18 | 1.11 | 1.25 | 0.10 |
| Hypotension | 0.89 | 0.69 | 1.16 |  | 0.81 | 0.48 | 1.34 |  | 0.98 | 0.84 | 1.14 | 0.69 |
| Myocardial infarction | 0.96 | 0.68 | 1.35 |  | 1.94 | 1.23 | 3.07 |  | 1.06 | 0.86 | 1.31 | 0.04 |
| Obesity | 1.42 | 1.19 | 1.69 |  | 0.78 | 0.52 | 1.18 |  | 1.08 | 0.94 | 1.23 | 0.01 |
| Other dysrhythmia | 1.00 | 0.88 | 1.13 |  | 1.01 | 0.81 | 1.26 |  | 1.22 | 1.13 | 1.32 | 0.01 |
| Psychosis | 0.84 | 0.76 | 0.92 |  | 0.93 | 0.79 | 1.09 |  | 1.08 | 1.02 | 1.14 | <0.01 |
| Hyperlipidemia | 0.95 | 0.87 | 1.03 |  | 0.90 | 0.78 | 1.04 |  | 0.90 | 0.85 | 0.95 | 0.58 |
| Pulmonary hypertension | 1.66 | 1.23 | 2.22 |  | 0.55 | 0.20 | 1.48 |  | 1.17 | 0.93 | 1.47 | 0.04 |
| Valvular heart disease | 1.78 | 1.52 | 2.08 |  | 0.71 | 0.46 | 1.10 |  | 1.41 | 1.25 | 1.58 | <0.01 |
| Sleep apnea | 1.14 | 0.88 | 1.46 |  | 0.71 | 0.41 | 1.21 |  | 1.16 | 0.99 | 1.36 | 0.22 |
| Smoking | 1.29 | 1.09 | 1.52 |  | 1.27 | 0.96 | 1.70 |  | 1.35 | 1.22 | 1.50 | 0.84 |
| Stroke | 1.07 | 0.89 | 1.29 |  | 1.04 | 0.74 | 1.48 |  | 1.38 | 1.23 | 1.54 | 0.04 |
| TIA | 0.95 | 0.73 | 1.25 |  | 0.99 | 0.61 | 1.62 |  | 0.74 | 0.61 | 0.88 | 0.21 |
| Angina | 1.24 | 1.03 | 1.50 |  | 1.21 | 0.87 | 1.70 |  | 1.13 | 1.00 | 1.29 | 0.73 |
| Renal insufficiency | 1.07 | 0.78 | 1.48 |  | 1.22 | 0.70 | 2.12 |  | 1.18 | 0.96 | 1.44 | 0.88 |
| Chronic renal disease | 1.49 | 1.29 | 1.73 |  | 1.57 | 1.22 | 2.04 |  | 1.63 | 1.48 | 1.79 | 0.62 |

\*EF $\geq 0.45$
†EF $< 0.45$
‡EF not recorded during 1 month before and 1 year from the date of first heart failure diagnosis
EF = Ejection fraction; COPD = chronic obstructive pulmonary disease; TIA = transient ischemic attack

**Figure 2.**
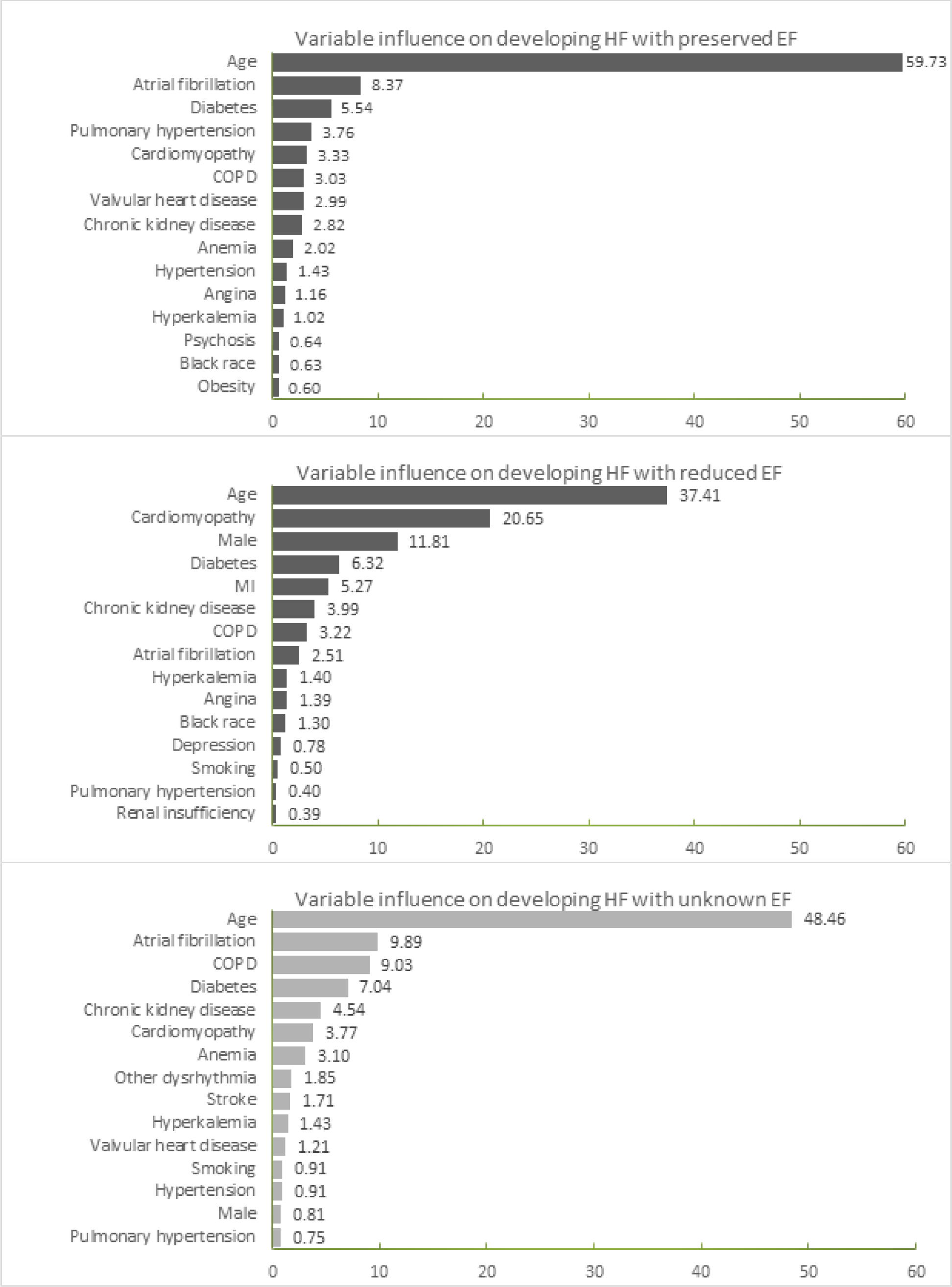
Variable Influence From GBM for HF With Reduced EF, Preserved EF or Uncertain EF.

#### Model performance

The c-index for the standard cox proportional hazards model was 0.73, and the performance of the competing risks model was identical. Discriminating accuracy of the GBM were also similar across heart failure types with c-index of 0.71, 0.70, and 0.75 for predicting heart failure with pEF, rEF and uncertain EF, respectively.

## DISCUSSION

In this population-based cohort study, we assessed the impact of various risk factors for different heart failure types and observed atrial fibrillation, obesity, pulmonary hypertension and valvular disease to be significantly associated with developing heart failure with pEF, while male gender, history of cardiomyopathy and myocardial infarction to be significantly associated with the risk of heart failure with rEF.

Heart failure incidence rate among individuals older than 65 years reported in the current study (20.9/1,000 person-years) is in line with observations from other population based cohorts (21/1,000 person-years). (1) Among commonly known risk factors for heart failure assessed in this study, several cardiovascular related conditions, diabetes, COPD, anemia, chronic kidney disease, smoking, and obesity were identified as potential predictors of heart failure, which is consistent with prior evidence. (8, 9) Dunlay et al. reported that the risk of incident heart failure increased by 3-fold with coronary disease and 2-fold with diabetes with the combined population attributable risk for coronary disease, hypertension, diabetes, obesity, and smoking of 52%. (9) Additionally, a cohort study involving elderly patients identified age, male gender, diabetes, hypertension, obesity, and myocardial infarction as important determinants of heart failure. (8)

Our findings on the differential effects of the risk factors across heart failure types are consistent with the findings from previous studies. In the Prevention of Renal and Vascular End-stage Disease (PREVEND) cohort study comparing the predictors of heart failure with rEF and pEF, atrial fibrillation was predominantly associated with heart failure with pEF, whereas myocardial infarction was predominantly associated with rEF. (12) In analyses of Framingham Heart Study participants, it was noted that higher BMI, smoking, and atrial fibrillation predicted incidence of pEF and male gender and history of cardiovascular disease predicted incidence of rEF. (10) The differences in the relationships between the underlying conditions and the subtypes of heart failure observed in our study provide additional support for the current consensus that heart failure with pEF and rEF may involve varying etiologies. (31)

Our understanding of the etiology of heart failure with pEF is evolving. A recent multi-cohort collaboration study demonstrated that currently established cardiovascular biomarkers including natriuretic peptides, high-sensitivity troponin, and C reactive protein were more strongly associated with heart failure with rEF than pEF, highlighting the potentially distinct pathophysiology of different heart failure subtypes and our limited knowledge regarding the underlying factors associated with the development of heart failure with pEF. (32) Based on the results from the GBM, we noted that aging plays the most significant role by far in incidence of pEF and other co-morbid conditions of atrial fibrillation, diabetes, and pulmonary hypertension, are also of high importance in development of pEF. These findings may be helpful in prioritizing populations for close monitoring of early signs of heart failure with pEF and targeting prevention efforts.

There are some unique strengths of our study. First, the total number of incident heart failure cases in our study is substantially larger (9,701) compared with the previous studies (374 in Brouwers et al. and 512 in Ho et al.), (10, 12) providing a higher statistical power for detecting the impact of individual risk factors on incidence of heart failure. Next, the mean age of our study population is 72 years, which is substantially higher than the previous studies (50 years in Brouwers et al. and 60 years in Ho et al.). (10, 12) Thus, our study focuses on elderly individuals with a high burden of co-morbid conditions, which is a population that is of great relevance with respect to heart failure incidence. However, there are also some limitations that deserve mention. First, for a large proportion of heart failure cases, we could not determine the type because of unavailability of EF results in our linked EHR system, and these may have been captured in other hospital systems where the Medicare patients received treatment. Heart failure cases with unmeasured LVEF may have represented a heterogeneous group of patients with rEF or pEF, which makes interpretation of results for this outcome difficult. Infrequent availability of EF values in investigations conducted using routine care heart failure populations has been noted in prior studies as well. (33, 34) This limitation can impact our study in two important ways: 1) our inability to classify nearly 60% of the cases into their respective heart failure subtype can compromise statistical power in detecting heterogeneous effects of the predictors across heart failure subtypes; 2) we grouped all patients with unavailable EF into a single category with the assumption that the EF is randomly missing in our EHR across classes of heart failure. However, if availability of EF in our data is dependent upon the subtype of heart failure, then the outcome misclassification resulting from this approach of classifying all patients with unavailable EF into a single group could lead to imperfect effect estimates of association between risk factors and development of rEF and pEF. Also, despite our efforts to include only patients without any heart failure by additionally excluding those with prior use of medications indicated for heart failure, misclassification of heart failure-free patients may still have occurred. Under-recording of certain non-severe chronic conditions in claims data is another limitation. For example, the negative associations of hyperlipidemia with heart failure may reflect potential discordance between the information available in claims data due to undercoding of chronic conditions among more severely ill patients. (35)

In conclusion, our findings of heterogeneous relationships between several important risk factors and heart failure types underline the potential differences in the etiology of heart failure with pEF and rEF. Amidst rising incidence and growing healthcare costs attributable to heart failure, it is critical to consider these potential differences between heart failure with pEF and rEF to develop effective surveillance and prevention strategies.

## Data Availability

The data that support the findings of this study are available from Partners Healthcare® but restrictions apply to the availability of these data, which were used under license for the current study, and so are not publicly available. Data are however available from the authors upon reasonable request and with permission of Partners Healthcare®.

